# Comparison of raw accelerometry data from ActiGraph, Apple Watch, Garmin, and Fitbit using a mechanical shaker table

**DOI:** 10.1101/2023.05.25.23290556

**Authors:** James W. White, Olivia Finnegan, Nick Tindall, Srihari Nelakuditi, David E. Brown, Russ Pate, Gregory J. Welk, Massimiliano de Zambotti, Rahul Ghosal, Yuan Wang, Sarah Burkart, Elizabeth L. Adams, Mvs Chandrashekhar, Bridget Armstrong, Michael W. Beets, R. Glenn Weaver

## Abstract

The purpose of this study was to evaluate the reliability and validity of the raw accelerometry output from research-grade and consumer wearable devices compared to accelerations produced by a mechanical shaker table. Raw accelerometry data from a total of 40 devices (i.e., n=10 ActiGraph wGT3X-BT, n=10 Apple Watch Series 7, n=10 Garmin Vivoactive 4S, and n=10 Fitbit Sense) were compared to the criterion accelerations produced by an orbital shaker table at speeds ranging from 0.6 Hz (4.4 milligravity-mg) to 3.2 Hz (124.7mg). For reliability testing, identical devices were oscillated at 0.6 and 3.2 Hz for 5 trials that lasted 2 minutes each. For validity testing, devices were oscillated for 1 trial across 7 speeds that lasted 2 minutes each. The intraclass correlation coefficient (ICC) was calculated to test inter-device reliability. Pearson product moment, Lin’s concordance correlation coefficient (CCC), absolute error, and mean bias were calculated to assess the validity between the raw estimates from the devices and the criterion metric. Estimates produced by the raw accelerometry data from Apple and ActiGraph were more reliable ICCs=0.99 and 0.97 than Garmin and Fitbit ICCs=0.88 and 0.88, respectively. Estimates from ActiGraph, Apple, and Fitbit devices exhibited excellent concordance with the criterion CCCs=0.88, 0.83, and 0.85, respectively, while estimates from Garmin exhibited moderate concordance CCC=0.59 based on the mean aggregation method. ActiGraph, Apple, and Fitbit produced similar absolute errors=16.9mg, 21.6mg, and 22.0mg, respectively, while Garmin produced higher absolute error=32.5mg compared to the criterion based on the mean aggregation method. ActiGraph produced the lowest mean bias 0.0mg (95%CI=-40.0, 41.0) based on the mean aggregation method. Raw accelerometry data collected from Apple and Fitbit are comparable to ActiGraph. However, raw accelerometry data from Garmin appears to be different. Future studies may be able to develop algorithms using device-agnostic methods for estimating physical activity from consumer wearables.

## Introduction

Over the past 20 years, the objective assessment of physical activity has improved due to the introduction of wearable monitors, such as accelerometers. Wearable monitors provide objective estimates of movement and overcome recall and desirability bias that may hamper self-reported measures of physical activity [1, 2]. Best practice recommendations for using accelerometers have shifted over the last decade from traditional activity counts (accelerations per a given epoch) [3] to using raw accelerometry data from accelerometers (i.e., x-, y-, and z-axis accelerometry data in ɡ’s typically collected multiple times per second) to estimate physical activity [4].

Consumer wearables (e.g., Apple Watch, Fitbit, Garmin) are increasingly popular measurement tools for assessing physical activity. Not only are these devices equipped with accelerometers to capture movement, but they are also unobtrusive and designed to be worn on the wrist, targeted for comfort and style, affordable for consumers, rechargeable, waterproof, and can be designed for children [5–8]. Technological advances allow consumer wearables to also frequently have extended battery life (i.e., up to 54 days) [9] and remote data capture and monitoring. For these reasons, there has been a multitude of measurement studies that have explored the validity of physical activity estimates produced by consumer wearables [10, 11].

However, these studies are limited because they rely on estimates of physical activity that are derived from proprietary algorithms developed by the companies that produce these devices (e.g., Apple, Garmin, Fitbit, etc.). This is a key limitation because these algorithms are unavailable for review by researchers [12]. The drawbacks of estimating physical activity based on proprietary algorithms are that it is unclear whether best practice recommendations were used to develop these algorithms, and the algorithms could be updated by these companies at any time unbeknownst to the user. Thus, estimates of physical activity collected from the same device across time may provide different estimates of activity due solely to changes in the underlying algorithms that produce these metrics.

An alternative, device-agnostic or monitor-independent approach may address these limitations by enabling data from any device to be processed using the same algorithm or processing methodology [13, 14]. A device-agnostic approach is a realistic possibility as consumer wearables have released application programming interfaces (API) that allow access to the raw accelerometry data (i.e., x, y, z axis readings collected by these devices [15]. This has the potential to increase the comparability of physical activity estimates across time and between different consumer wearables and research-grade devices.

A necessary first step to applying a device-agnostic approach to raw accelerometry data collected by consumer wearables is to conduct calibration studies that explore the reliability and validity of the underlying acceleration output produced by these devices [16]. This testing will provide insight into the reliability and validity of the raw acceleration output from consumer wearables in a controlled environment, prior to evaluating how human variation impacts the raw acceleration estimates from these devices [16]. Therefore, this study will evaluate the between-device reliability and validity of the raw acceleration output from research-grade and consumer wearable devices, compared to accelerations produced by a mechanical shaker table at various speeds as the criterion measure. It is important to include research-grade devices in this study because it allows us to evaluate if the raw accelerometry estimates from consumer wearables are comparable to the raw accelerometry estimates of research-grade devices when compared to more direct estimates of acceleration from a mechanical shaker table. While studies have previously examined the ActiGraph with this methodology [17, 18], this is the first study that we are aware of to report shaker table outcomes with consumer-grade devices.

## Methods

Raw accelerometry data from a total of 40 devices were evaluated in this study. The research-grade devices included n=10 ActiGraph wGT3X-BT (ActiGraph; ActiGraph LLC Pensacola, FL). The consumer wearable devices included n=10 Apple Watch Series 7 (Apple; Apple Technology Company, Cupertino, CA), n=10 Garmin Viovactive 4S (Garmin; Garmin Ltd., Olathe, KS), and n=10 Fitbit Sense (Fitbit; Google LLC, San Francisco, CA). Inter-device reliability and validity of raw accelerations for all devices were tested, with accelerations produced by a mechanical shaker table (Scientific Industries, Bohemia, NY; Mini-300 Orbital-Genie, Model 1500) as the criterion. Each device was securely mounted directly to the twin ratcheting clamps of a mechanical shaker table (Fig 1) that produces controlled oscillations at frequencies between approximately *f_shaker_*=0.6 and 5 Hertz (Hz). We converted *f_shaker_* in Hz to acceleration using the expression for centripetal acceleration, *a_orbital_* = *v*^2^/*r_orbital_* [19], where *r_orbital_* is the radius of rotation for the orbital shaker *r_orbital_*. From the manual for this particular shaker (supplementary https://cdn.shopify.com/s/files/1/0489/6990/8374/files/SI-M1600_Manual.pdf?v=1617998279), the specified diameter of the orbit is 2*r_orbital_*=1.9cm and the rotational speed is given by *v* = 2π *r_orbital_f_shaker_*, since for each complete cycle of 2π radians, the table traverses a distance of circumference 2π*r_orbital_* in time 1/*f_shaker_*. In other words:

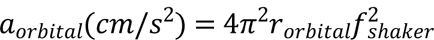

to convert this acceleration to units of earth’s gravity (g’s), divide *a_orbital_* by 9.81cm/s^2^.

**Figure 1.**
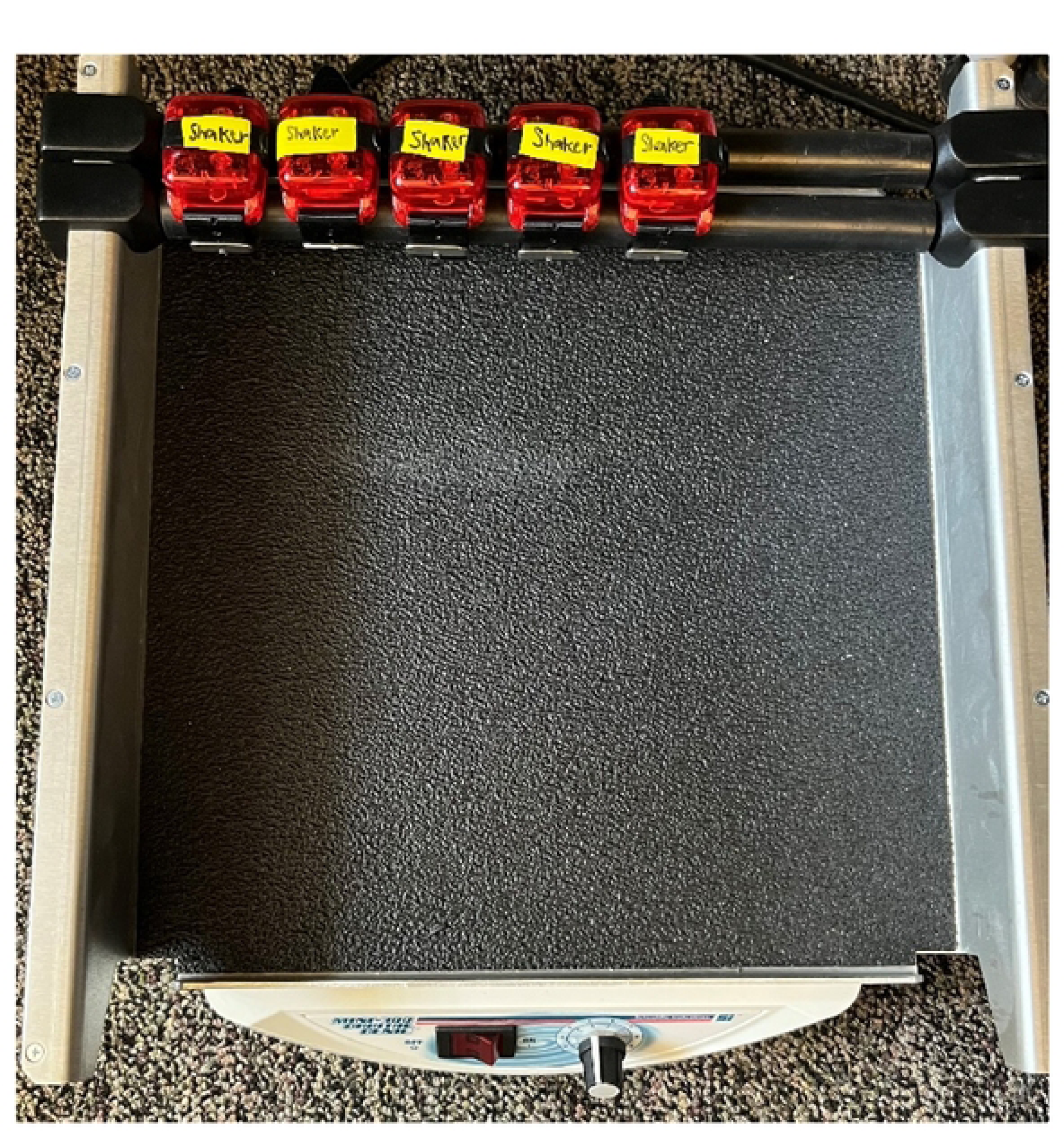
Orbital mechanical shaker used for shaker testing.

A total of five devices were placed on the shaker table at once. Serial number/device ID and position of devices (numbered 1 to 5 from left to right) were recorded for all devices. Prior to each trial, the shaker table was placed on a level surface (i.e., floor); time from each device was recorded at the second level.

### Device software

ActiGraphs were initialized to provide output from each directional axis using ActiLife software (version 6.13.4; ActiGraph LLC, Pensacola, FL). Garmin devices were initialized, and data were recorded in RawLogger (version 1.0.20211201a) and exported through Garmin Connect software^TM^. Apple devices were initialized, and data were recorded in SensorLog (version 5.2) and exported into comma-separated values (CSV) files via Health Auto Export (version 6.3). RawLogger and SensorLog are user-written apps that leverage the device-specific Application Programming Interface (API) to collect the underlying sensor data on the respective devices. RawLogger is available for download through the Connect IQ^TM^ store on the Garmin Connect^TM^ app, and SensorLog and Health Auto Export are available for download through the App Store. The research team developed a custom Fitbit app (Slog) leveraging the Fitbit API for the same purpose, and Fitbit devices were initialized, and data were recorded and exported through this app. The GitHub code for the custom Fitbit app is available at https://github.com/ntindallUSC/Slog/tree/main. Sampling frequencies from 25 Hz to 100 Hz were recorded based on the capabilities of the ActiGraph (100 Hz), Apple (100 Hz), Garmin (25 Hz), and Fitbit (50 Hz).

### Reliability testing

Reliability testing included five identical devices mounted side-by-side (e.g., 5 ActiGraph devices) positioned 1-5. Each device was tested for five 2-minute trials at 0.6 Hz and 3.2 Hz for a total of 10 trials until all devices were tested. A 15-second rest period took place at the beginning and end of each trial. Thus, it took ten minutes and 30 seconds to test 5 devices at one speed. The time of the 15-second rest periods and the trial start and end time were recorded based on device time. A minimum of 20 trials were conducted for each device brand, totaling 80 trials. Trials with missing data due to device malfunction: Apple (n=20) and Fitbit (n=10) were repeated to ensure that raw acceleration data from all devices could be analyzed; no trials had to be repeated for ActiGraph and Garmin devices.

### Validity testing

For validity testing, five identical devices were mounted side-by-side until all devices were run through the validity trials. The trials lasted 14 minutes and 30 seconds. Consistent with past validation studies [18, 20], each trial began with a 15-second rest period (i.e., no movement) followed by a standardized series of oscillations at seven frequencies (i.e., 3.2 Hz, 2.8 Hz, 2.4 Hz, 1.9 Hz, 1.5 Hz, 1.0 Hz, 0.6 Hz) lasting two minutes each. These frequencies were chosen because they are consistent with human movement ranging from 1.5 to 16 mph [21]. The start and stop times were noted at each frequency for both research-grade and consumer wearable devices. Each trial ended with another 15-second rest period. A minimum of 2 trials were conducted for each device brand, totaling 8 trials. Trials/devices with missing data due to device malfunction: Apple (n=4) and Fitbit (n=1) or shaker table malfunction (n=1) were repeated to minimize missing data; no trials had to be repeated for ActiGraph or Garmin devices. Following all testing, raw acceleration data for both research-grade and consumer wearable devices were downloaded and converted to a CSV file using ActiLife software and the device-specific user-written apps, respectively.

### Data processing

Raw acceleration data from all devices (i.e., ActiGraph, Apple, Garmin, and Fitbit) were extracted from the middle minute of each 2-minute oscillation frequency. Consistent with past research, Euclidean Norm Minus One (ENMO) was calculated [4, 5]. All values were multiplied by 1000 (milligravity-mg) to be consistent with published intensity thresholds based on the GGIR package for accelerometry in R statistical software [22]. Data were aggregated to the second level by extracting the mean and root mean square (RMS) value for each second for all devices for ENMO. Both mean and RMS were calculated as both methods have been calculated previously, which suggests that there is no consensus on how raw accelerometry data should be aggregated [18, 20, 23].

### Correlation coefficients

To test reliability, a single, absolute intraclass correlation coefficient (ICC) was calculated for all devices. ICC values less than 0.50 were defined as poor reliability, between 0.50 and 0.75 as moderate reliability, between 0.75 and 0.90 as good reliability, and greater than 0.90 as excellent reliability [24]. Prior to statistical analyses for validity testing, descriptive means and standard deviations for the mean and RMS were calculated across devices for each speed ranging from 0.6 to 3.2 Hz. For the validity testing, Pearson product moment *(r)* and Lin’s concordance correlation coefficient (CCC) were calculated to assess correlation and agreement of raw acceleration data from ActiGraph and consumer wearable devices compared to the criterion (i.e., acceleration from the shaker table) [25]. Pearson product moment interpretations were defined based on Dancey and Reidy [26], and Lin’s concordance correlation coefficient was defined similarly based on recommendations from Altman (1991), with coefficients less than 0.20 as poor and greater than 0.80 as excellent [27].

### Discrepancy analyses

An absolute error was calculated to assess the magnitude of the error between the criterion metrics and the raw acceleration data from ActiGraph and consumer wearable devices. The mean bias was also calculated to assess whether the raw acceleration output from ActiGraph and consumer wearable devices over-or underestimated acceleration output compared to the criterion metric. Raw acceleration data from one ActiGraph (ID=210) was eliminated because the device was faulty and provided implausible acceleration values (all ENMO values were below 0). Thus, there were (N=3,780) observations for ActiGraph, whereas Apple and Garmin devices contributed (N=4,200) observations. Missing data were present across all Fitbit devices except two, which contributed to (N=3,975) observations for Fitbit.

## Results

For reliability, ICCs (95% confidence intervals) are presented for the raw acceleration data from all devices for both aggregation methods (i.e., mean and RMS) for all devices in Table 1. The ICCs for ActiGraph were 0.97 (0.92, 0.99) and 0.97 (0.93, 0.98) for the mean and RMS aggregation methods, respectively. The ICCs for Apple were 0.99 (0.99, 0.99) and 0.99 (0.99, 1.00) for the mean and RMS, respectively. The ICCs for Garmin were 0.88 (0.82, 0.92) and 0.90 (0.85, 0.93) for the mean and RMS aggregation methods, respectively. The ICCs for Fitbit were 0.88 (0.86, 0.89) and 0.87 (0.85, 0.88) for the mean and RMS aggregation methods, respectively.

**Table 1.**
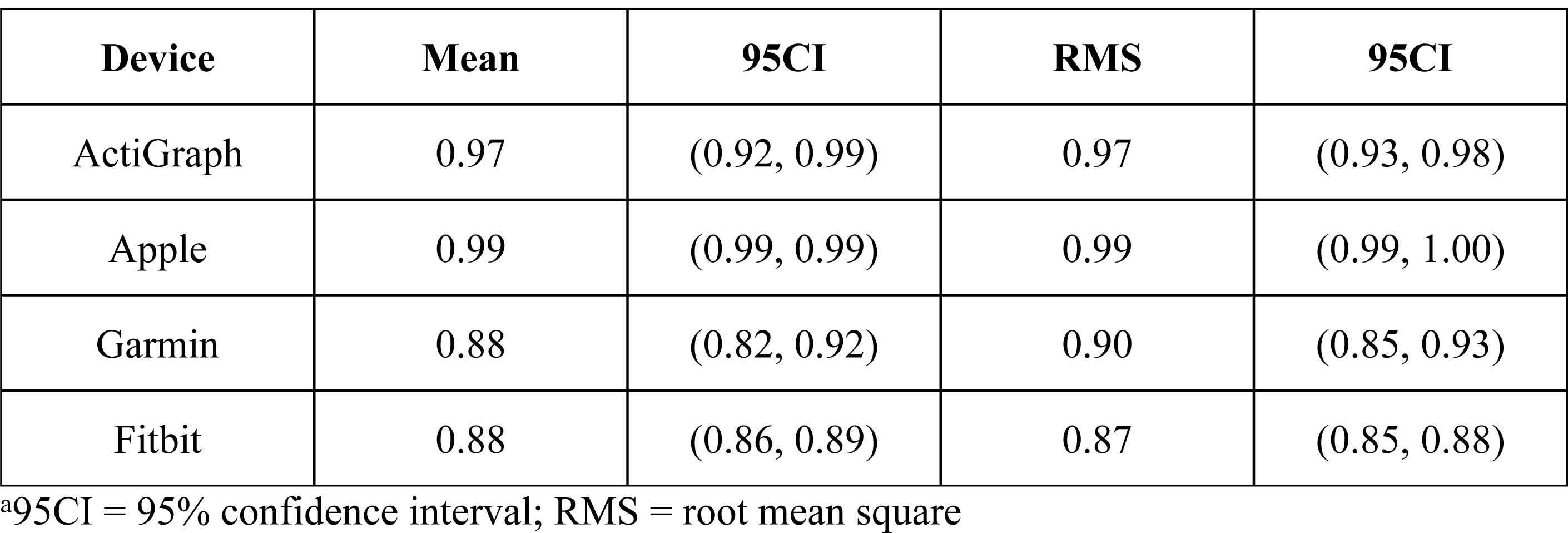
Summary of Intraclass Correlation Coefficients for All Devices Aggregated based on the Mean and Root Mean Square.

| Device | Mean | 95CI | RMS | 95CI |
| --- | --- | --- | --- | --- |
| ActiGraph | 0.97 | (0.92, 0.99) | 0.97 | (0.93, 0.98) |
| Apple | 0.99 | (0.99, 0.99) | 0.99 | (0.99, 1.00) |
| Garmin | 0.88 | (0.82, 0.92) | 0.90 | (0.85, 0.93) |
| Fitbit | 0.88 | (0.86, 0.89) | 0.87 | (0.85, 0.88) |

<sup>a</sup>95CI = 95% confidence interval; RMS = root mean square

For validity, a summary table of outcomes based on the raw acceleration data from all devices is presented in Table 2. Fig 2 shows the concordance of the raw acceleration data from all devices compared to the criterion metric. Fig 3 shows the absolute error of the raw acceleration data from all devices compared to the criterion metric. Fig 4 shows the mean bias of the raw acceleration data from all devices compared to the criterion metric.

**Fig 2.**
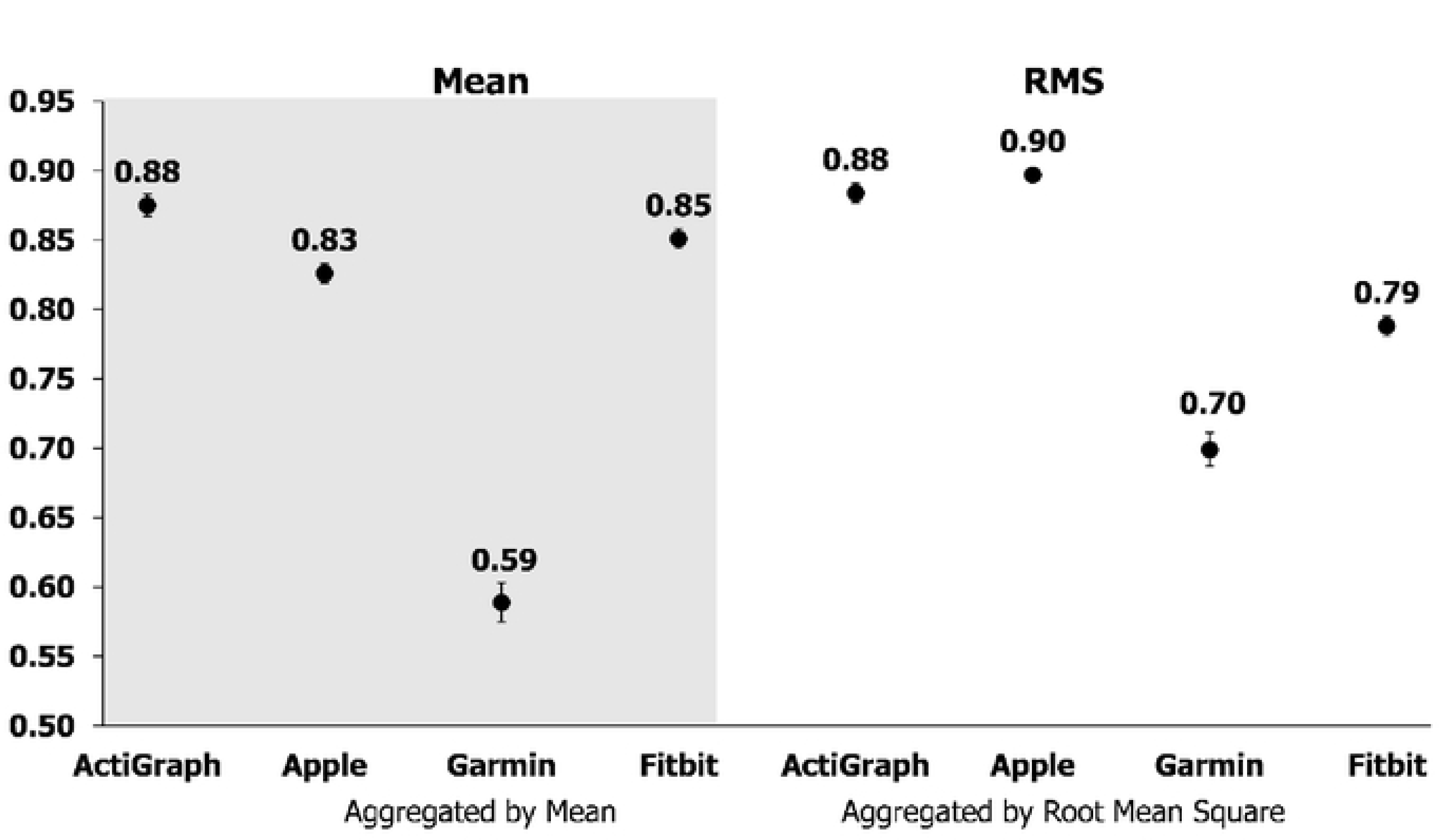
Lin’s Concordance Correlation Coefficient of the Raw Acceleration Data from all Devices Compared to the Accelerations Produced by a Mechanical Shaker Table.

**Fig 3.**
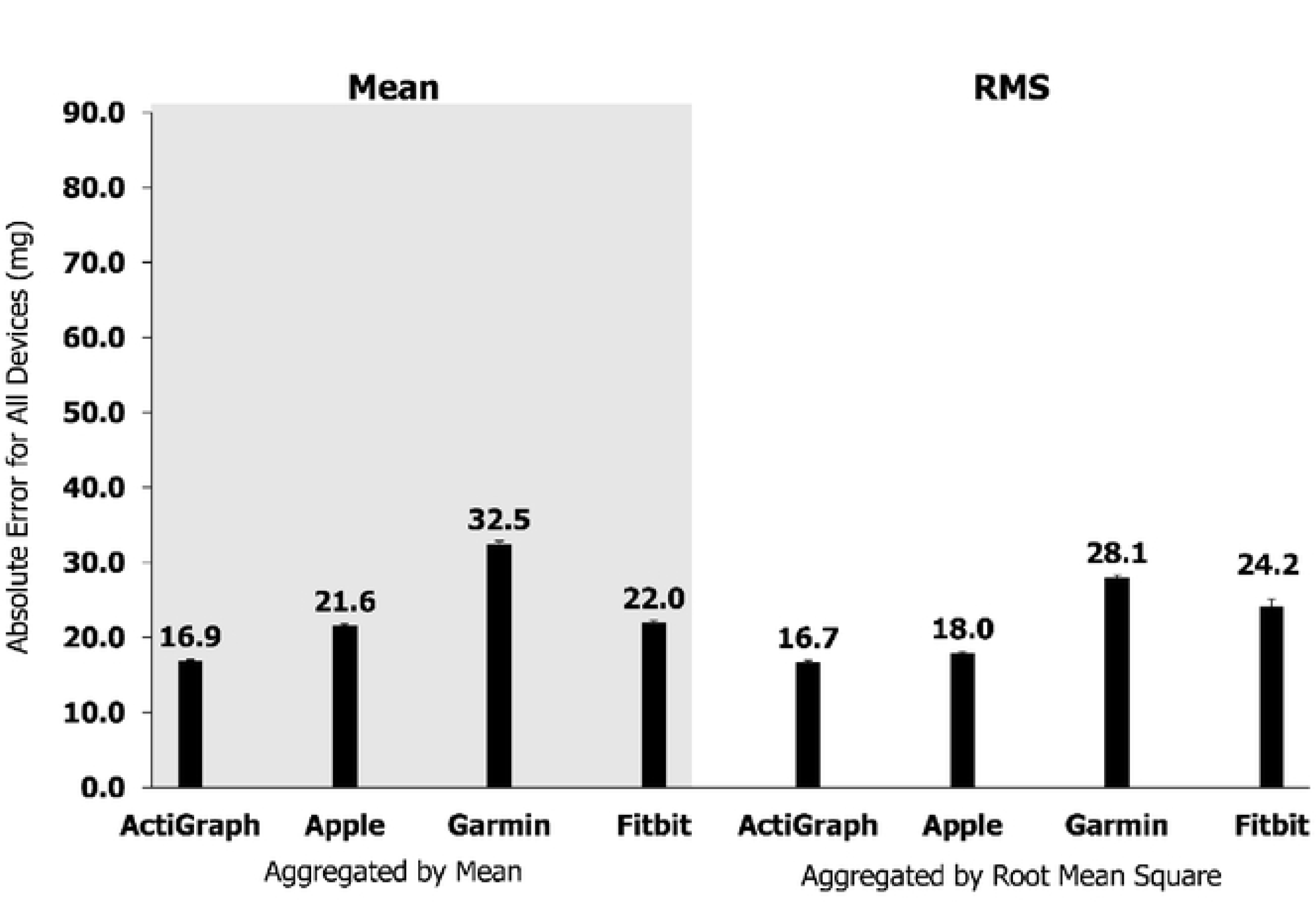
Absolute Error of the Raw Acceleration Data from all Devices Compared to the Accelerations Produced by a Mechanical Shaker Table.

**Fig 4.**
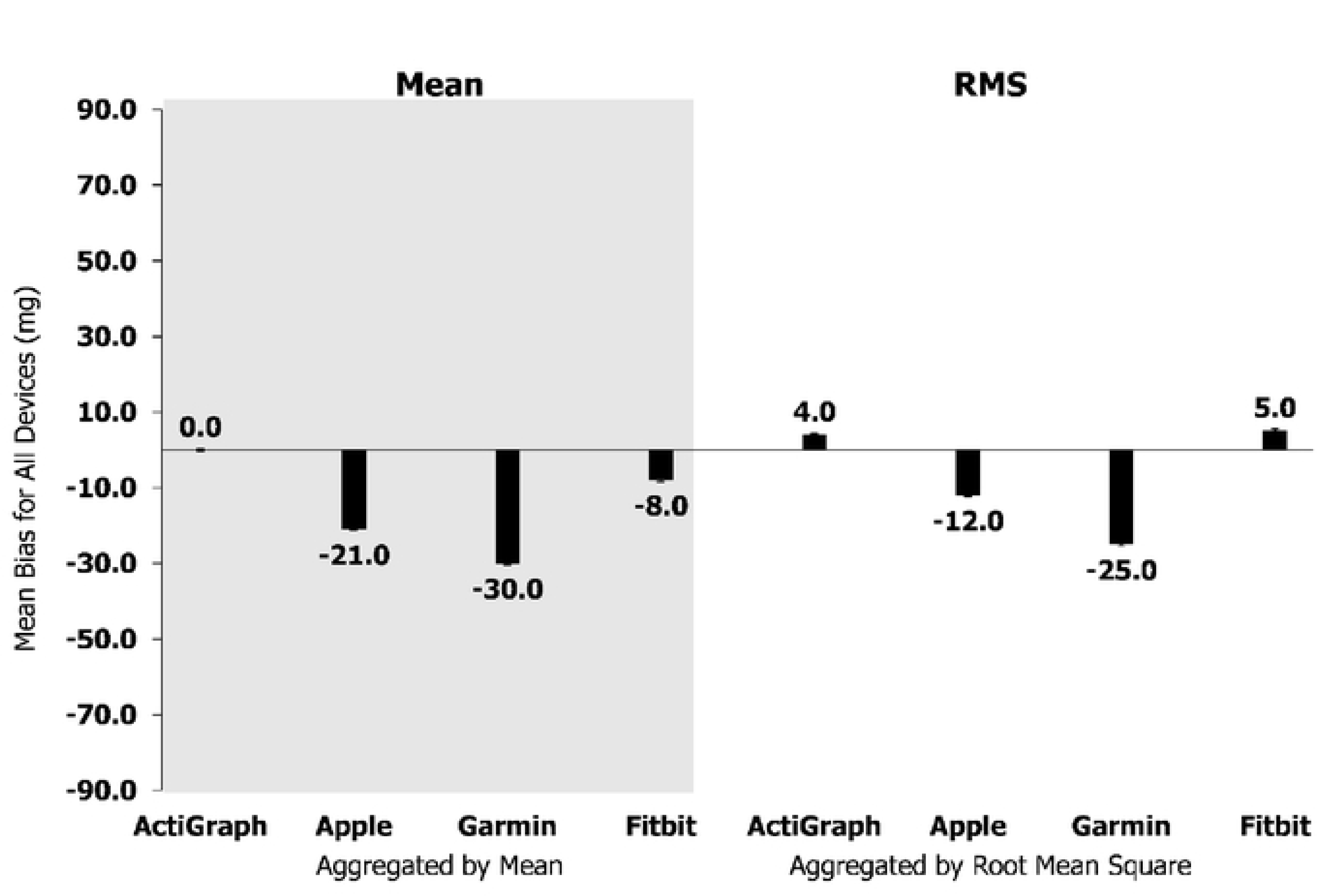
Mean Bias of the Raw Acceleration Data from all Devices Compared to the Accelerations Produced by a Mechanical Shaker Table.

**Table 2.** Summary of Validity Outcomes for All Devices Aggregated based on the Mean and Root Mean Square.

|  | Devices | ActiGraph | Apple | Garmin | Fitbit |
| --- | --- | --- | --- | --- | --- |
| **Mean** | Observations | 3,780 | 4,200 | 4,200 | 3,975 |
|  | Mean (mg) | 54.4 | 32.7 | 23.8 | 46.1 |
|  | SD (mg) | 41.5 | 41.0 | 34.1 | 57.4 |
|  | Pearson's r | 0.88 | 0.94 | 0.79 | 0.91 |
| **Root Mean Square** | Observations | 3,780 | 4,200 | 4,200 | 3,975 |
|  | Mean (mg) | 58.1 | 41.8 | 29.0 | 58.8 |
|  | SD (mg) | 45.0 | 48.9 | 37.9 | 71.8 |
|  | Pearson's r | 0.89 | 0.94 | 0.84 | 0.92 |

<sup>a</sup>SD = standard deviation; mg = milligravity

Pearson product moment correlations between raw accelerometry estimates for ActiGraph and the criterion metric were *r=*0.88 and *r=*0.89 for the mean and RMS aggregation methods, respectively. CCCs (95% confidence intervals) when compared to the shaker table were *r_c_*=0.88 (0.87, 0.80) and *r_c_*=0.88 (0.88, 0.89) for the mean and RMS aggregation methods, respectively. Mean bias (95% confidence intervals) was 0.0mg (−40.0, 41.0) and 4.0mg (−36.0, 44.0), and absolute error was 16.9mg and 16.7mg for the mean and RMS aggregation methods, respectively.

Pearson product moment correlations between raw accelerometry estimates for Apple and the criterion metric were *r=*0.94 and *r=*0.94 for the mean and RMS aggregation methods, respectively. CCCs when compared to the shaker table were *r_c_*=0.83 (0.82, 0.83) and *r_c_*=0.90 (0.89, 0.90) for the mean and RMS aggregation methods, respectively. Mean bias (95% confidence intervals) was −21.0mg (−50.0, 7.0) and −12.0mg (−45.0, 21.0), and absolute error was 21.6mg and 18.0mg for the mean and RMS aggregation methods, respectively.

Pearson product moment correlations between raw accelerometry estimates for Garmin and the criterion metric were *r=*0.79 and *r=*0.84 for the mean and RMS aggregation methods, respectively. CCCs when compared to the shaker table were *r_c_*=0.59 (0.58, 0.60) and *r_c_*=0.70 (0.69, 0.71) for the mean and RMS aggregation methods, respectively. Mean bias (95% confidence intervals) was −30.0mg (−80.0, 19.0) and −25.0mg (−69.0, 19.0), and absolute error was 32.5mg and 28.1mg for the mean and RMS aggregation methods, respectively.

Pearson product moment correlations between raw accelerometry estimates for Fitbit and the criterion metric were *r=*0.91 and *r=*0.92 for the mean and RMS aggregation methods, respectively. CCCs when compared to the shaker table were *r_c_*=0.85 (0.84, 0.86) and *r_c_*=0.79 (0.78,0.80) for the mean and RMS aggregation methods, respectively. Mean bias (95% confidence intervals) was −8.0mg and 5.0mg, and absolute error was 22.0mg and 24.2mg for the mean and RMS aggregation methods, respectively.

## Discussion

The aim of this study was to evaluate the between-device reliability and validity of the raw acceleration output from research-grade (i.e., ActiGraph wGT3X-BT) and consumer wearable devices (i.e., Apple Watch Series 7, Garmin Vivoactive 4S, and Fitbit Sense) compared to accelerations produced by a mechanical shaker table. The raw acceleration data collected from all devices exhibited good-to-excellent between-device reliability based on the mean and RMS aggregation methods. For validity, the raw acceleration data from all devices exhibited a strong positive correlation to the criterion metric with moderate-to-excellent concordance no matter the aggregation method. Except for Garmin, the raw acceleration data collected from consumer wearables demonstrated absolute errors that were consistent with ActiGraph. Moreover, the raw acceleration data collected from consumer wearables underestimated acceleration output to a greater degree than ActiGraph when compared to the accelerations produced by the mechanical shaker table. Overall, the raw acceleration data for all devices differed when data were aggregated based on the mean and RMS for each second, with values generally being more reliable and accurate based on the RMS aggregation method.

A key finding of this study is that the reliability for Apple, Garmin, and Fitbit was similar to ActiGraph. In fact, consumer wearables exhibited moderate-to-excellent ICC values, with Apple demonstrating nearly perfect reliability with an ICC of 0.99. These findings are similar to other studies evaluating the between-device reliability of research-grade devices using a mechanical shaker table. For instance, Powell et al. [28] reported an ICC of 0.99 between 23 RT3 accelerometers and Santos-Lozano et al. [17] reported an ICC of 0.97 between 10 ActiGraph GT3X accelerometers. More recently, studies have explored within-device reliability of various accelerometers and have reported ICCs ranging from 0.77 to 1.00 [29, 30]. Thus, ICCs based on the raw acceleration data collected from consumer wearables in the present study support their use as a reliable tool to assess physical activity.

In the present study, it is also important to note that raw accelerometry estimates collected from Apple and Fitbit exhibited correlation and concordance with the criterion metric that was consistent with ActiGraph. On the other hand, raw acceleration data collected from Garmin exhibited less correlation and concordance with the criterion metric than ActiGraph. Our findings for Apple and Fitbit correlation are more consistent with a previous study that reported an excellent Pearson correlation (*r*=0.97) for accelerations produced by GENEA accelerometers and a mechanical shaker table [23]. These findings suggest that raw acceleration data from Apple and Fitbit may produce comparable estimates of activity than raw acceleration data from ActiGraph. More information is needed to determine whether the raw acceleration data from Garmin could be used to accurately estimate physical activity. These findings could be due to the hardware differences between devices. For example, the dynamic accelerometer range of the ActiGraph is ±8g [31], while the default accelerometer range for Fitbit is ±4g [32]. The dynamic accelerometer range is an estimate of the greatest amount of acceleration that a device can accurately assess, and thus the relatively smaller accelerometer range of Garmin and Fitbit compared to ActiGraph could have led to more error in Garmin and Fitbit raw accelerometry estimates at greater frequencies (S Fig 1 and 2). Differences in the raw acceleration output collected from ActiGraph and the consumer wearables could also be due to the post-processing of the raw data, which has been described previously [18].

Further evidence revealed that, compared to the criterion metric, raw acceleration estimates from Apple and Fitbit exhibited absolute errors similar to the raw acceleration estimates from ActiGraph, while raw acceleration estimates from Garmin exhibited larger absolute errors relative to the raw acceleration estimates from ActiGraph. It is also important to note that raw acceleration data from Apple and Garmin underestimated acceleration output by more than 20mg and 30mg, respectively, compared to raw acceleration estimates from ActiGraph. This evidence is concerning for Garmin, considering that published intensity thresholds derived from ActiGraph worn on the non-dominant wrist indicates that sedentary thresholds for children (7-11yrs) are under 35.6mg [33, 34]. Based on these intensity thresholds, it would be difficult to distinguish between sedentary and light intensity thresholds for children using raw acceleration output from Garmin. This may suggest that we need to move away from cut-points, especially since a device-agnostic approach may allow for increased comparability of physical activity estimates across time and between consumer wearables and research-grade devices. However, more work is needed, specifically with Garmin. A device-agnostic approach using raw accelerometry data from Garmin could lead to different estimates of activity because the raw accelerometry output is different from ActiGraph, Apple, and Fitbit.

Overall, the findings suggest that the raw acceleration output from Apple and Fitbit is comparable to the raw acceleration output from ActiGraph. However, limitations with accelerometry are well-documented for distinguishing between sedentary and light activity. For instance, a study using 2-regression models to estimate energy expenditure derived from ActiGraph counts observed mean absolute percent error values that ranged from 32.5% to 39.4% and 14.5% to 42.9% for sedentary and light activities, respectively, in children 7-13yrs [35]. A similar study reported that research-grade accelerometers (i.e., ActiGraph, Actical, and AMP-331) tended to overestimate sedentary and light activities in adults [36]. Though most of the evidence on the associations of objectively assessed sedentary behavior and health is based on accelerometers that infer sedentary time from a lack of movement, this can lead to misclassification of low-movement, non-sedentary behaviors as sedentary behaviors [37]. The absolute errors of ActiGraph, Apple, and Fitbit (∼20mg) compared to the criterion metrics suggest that the relatively small window for sedentary behavior (under 35.6mg) may pose an issue for estimating physical activity outcomes from accelerometry [22]. Therefore, additional metrics (i.e., heart rate) may need to be combined with accelerometry to improve estimates of these outcomes. An advantage of consumer wearables is their ability to collect acceleration and heart rate data simultaneously. Thus, it may be possible to leverage the raw acceleration and heart rate data from consumer wearables (i.e., Apple and Fitbit) to overcome limitations with accelerometry alone for estimating physical activity outcomes.

There were several strengths of the present study. The first strength is that accelerations produced by a mechanical shaker table served as the criterion to assess the reliability and validity of accelerations produced by various accelerometers. This method allowed for a highly controlled, repeatable evaluation of underlying accelerations produced by various accelerometers shaken in orbital motion at known frequencies. Another strength is that the raw accelerations from devices were evaluated, allowing for between-monitor comparisons of accelerations through elimination of proprietary signal processing that has traditionally been used to derive activity counts from research-grade devices [18]. Additionally, this study evaluated the raw accelerations from consumer wearables, addressing concerns about the proprietary signal processing of these devices [38]. By evaluating the raw accelerations for both research-grade and consumer wearable devices, we were able to compare acceleration estimates from the devices based on the same metric (mg). Lastly, we calculated Lin’s CCC, absolute error, and mean bias to assess the agreement of the raw accelerometry data from research-grade and consumer wearable devices compared to accelerations produced by the shaker table. This allowed us to evaluate the agreement of the accelerations between proxy and criterion, the overall error of the raw acceleration estimates, and the direction of the average error of the raw acceleration estimates from all devices, whereas other studies used Pearson correlation to assess validity [20, 23].

Pearson correlation merely measures the covariance between two variables, not the agreement or error. Using these statistics, we were also able to compare the validity metrics produced by the raw acceleration estimates from consumer wearables to the validity metrics produced by the raw acceleration estimates from a research-grade device. This provided preliminary evidence for using the raw acceleration output of consumer wearables to estimate physical activity outcomes. However, the raw acceleration output from consumer wearables needs to be evaluated in settings that resemble free-living activities for children.

The limitations of the present study also need to be acknowledged. One limitation may be the technological advances that have occurred in the consumer wearables evaluated during the project. For instance, the Apple Watch Series 8 was released during the project. However, most of the technological advancements between the Apple Watch Series 7 and the Apple Watch Series 8 are centered on the dual-core processor and the addition of a temperature sensor [39], and thus may not impact accelerometer estimates between devices. Yet, information about the hardware of accelerometers used in consumer wearable devices is largely proprietary. Another limitation may be the post-processing of the raw acceleration data for all devices [18]. The post-processing of the raw acceleration data for all devices is proprietary, so the acceleration data is not truly raw. It is also unclear why missing data were present across all Fitbit devices except two. This may have been due to software malfunction with the custom Fitbit app (Slog) that was used to leverage the Fitbit Application Programming Interface.

## Conclusions

Findings from this study suggest that raw accelerometry data from Apple, Garmin, and Fitbit are reliable and provide estimates of raw accelerometry that are similar to ActiGraph. Additionally, raw accelerometry estimates for Apple and Fitbit are comparable to raw accelerometry estimates from ActiGraph, while raw accelerometry estimates from Garmin differ from estimates from ActiGraph. Yet, limitations with accelerometry are well-documented for distinguishing between sedentary and light activity. Consumer wearables’ ability to capture both accelerometry and heart rate could improve estimates of activity, especially sedentary and light activity. Future studies should explore using a device-agnostic approach for estimating physical activity from raw accelerometry data produced by Apple and Fitbit in settings that resemble free-living activities for children.

## Data Availability

Data cannot be shared publicly because of the institutional review board policies of the lead author. The data underlying the results presented in the study are available upon reasonable request.

## Acknowledgements

None.

## Supporting information

**S1 Fig. Absolute Error of the Raw Acceleration Data from all Devices by Speed Compared to the Accelerations Produced by a Mechanical Shaker Table.**

**S2 Fig. Mean Bias of the Raw Acceleration Data from all Devices by Speed Compared to the Accelerations Produced by a Mechanical Shaker Table.**

